# Prevalence of malnutrition among children at primary cleft surgery: A cross-sectional analysis of a global database

**DOI:** 10.1101/2021.01.20.21250177

**Authors:** Barbara Delage, Marko Kerac, Erin Stieber, Pamela Sheeran

## Abstract

**Background:** Orofacial clefts are common birth defects requiring prompt feeding support and timely surgery. Little information exists about the impact of inadequate care provision in poor-resource settings. We aimed to estimate the burden of malnutrition in children from 101 low- and middle-income countries (LMICs) using cleft surgery records collected by one cleft NGO.

**Methods:** We conducted a cross-sectional study using anonymised records of children ≤5 years who underwent cleft surgery between 2008 and 2018. The data included birth date, gender, weight at surgery, ethnicity, country of origin, and date of primary surgery and was analysed using descriptive statistics. The prevalence of malnutrition was derived from the generation of weight-for-age z scores and described in relation to cleft type, gender, and ethnicity/geography. For purpose of comparison, the most recent prevalence estimates for underweight in children under-5 were extracted from publicly available national surveys.

**Findings:** The analysis included 602,568 children. The overall prevalence of underweight at the time of primary cleft surgery was 28**·**6% **–** a figure well above the global underweight prevalence in under-5 children without cleft estimated at about 13**·**5%. The prevalence of underweight varied with the age at primary surgery and the type of cleft, as well as with gender, ethnicity, and region of origin, and was positively correlated with country-specific estimates of underweight prevalence in children without cleft.

**Interpretation:** Our findings support the need for advancing universal health coverage with special efforts to increase timely nutrition care and access to surgery for the most disadvantaged children.

**Funding:** None.

**Research in context:** *Evidence before this study:* Regardless of the setting, infants born with an orofacial cleft have a heightened risk of failure to thrive (FTT), especially when their ability to suck and swallow is compromised.(1-3) Timely identification of feeding problems and appropriate nutrition support are essential to ensure healthy child development.(4-6) Limited access to (specialist) care in LMICs increases the risk of FTT in babies with unrepaired cleft, yet limited research has described the extent of the problem in these settings. We searched Medline and Google Scholar up to April 2020 for studies that estimated the scale of malnutrition in children with cleft born in limited-resource settings. A 2019 systematic review of the literature identified seven cross-sectional or case-control studies conducted in LMICs (three in Africa(7-9), three in Latin America(10-12), one in Iran(13)).(14) We excluded one study in Brazil(10) which did not estimate undernutrition and found one additional cross-sectional study from South Africa.(15) Overall, seven hospital-based studies published between 1999 and 2017 included a total of 2,300 children <5 years old. They all provided evidence of malnutrition in this population, yet none was designed to give a global prevalence estimate.

*Added value of this study:* This study is the first that attempted to provide a global prevalence estimate of malnutrition in children with unrepaired cleft in LMICs. Using pre-surgery clinical records from over 600,000 of patients operated by Smile Train’s global partners, we identified underweight in 28·6% of children ≤5 years. Country-specific figures ranging from 6·9% in Kazakhstan to 48·2% in Chad were above national statistics on the prevalence of underweight in children in the general populations. Cleft epidemiology contributes to variations in malnutrition rates across LMIC settings but do not explain health disparities between children with cleft and those without cleft within countries.

*Implications of all the available evidence:* There is an urgent need to identify and/or address the barriers that prevent children with cleft from receiving immediate feeding and nutritional support and timely reparative surgery. Current health services and nutrition programmes in LMICs should consider opportunities to help meet the health care needs of these children. Poor early-life nutrition has well-documented detrimental consequences on child physical, functional, and cognitive development. Accordingly, a higher prevalence of malnutrition in children born with a cleft means that this population likely experiences higher rates of morbidity and mortality – even if they are eventually operated.

## Introduction

There is a great global disparity in the care of children born with orofacial cleft, a curable condition that requires immediate attention, age-appropriate surgical repair, and comprehensive care. In high-income countries, perinatal nursing care ensures an early diagnosis and the monitoring of any feeding and/or breathing issues that could interfere with growth and development, especially in infants with a cleft palate. Cleft care involves a multidisciplinary team of specialist health workers like lactation specialists, nutritionists, paediatricians, anaesthesiologists, cleft surgeons, ENT specialists, dentists, orthodontists, and speech therapists.(16) In contrast, identification, specialist attention and resources for cleft repair are lacking in many low- and middle-income countries (LMICs). In several LMICs, the coverage of essential yet basic maternal and child health services is low and less than half of all births are assisted by a skilled health attendant.(17) In addition, less than 5% of South Asians and Sub-Saharan Africans have access to timely, safe, and affordable surgical care.(18) In these circumstances, many newborns with cleft are not identified and/or operated and the impact of cleft on their nutritional status and survival is presumably significant, especially in contexts of widespread malnutrition and high disease burden.

While a handful of small-size studies have reported high rates of malnutrition among children with cleft in low-resource settings(14), the global burden of malnutrition among unrepaired children with cleft in LMICs remains unknown. This study aimed to estimate the prevalence of underweight using clinical data collected by one large nongovernmental organization subsidizing cleft surgery and comprehensive care services across the developing world.(19)

## Methods

### Study design and participants

This cross-sectional study included records of individuals who underwent primary surgery for orofacial clefts in Smile Train-sponsored partner facilities in LMICs. Only information uploaded by local cleft care providers into Smile Train’s online medical database between July 1, 2008 and June 30, 2018 has been retrieved (N=868,854 entries). The study was restricted to patients aged ≤5 years at the time of surgery (N=638,988 entries). Reasons for this included the fact that (i) orofacial clefts can complicate the feeding of newborns and immediately place them at risk of malnutrition, (ii) WHO growth standards are available to assess the nutrition status of children ≤5 years old, and (iii) country-specific estimates on malnutrition in children aged ≤5 years are accessible from the Demographic and Health Survey (DHS) program for comparison.

### Variables

Anonymised records of patients included date of birth, gender, weight at surgery, ethnicity, country of origin, cleft type, and date of cleft operation. Age at surgery was calculated using date of birth and date of operation and any erroneous values (≤0 years) or values >5 years were excluded. Out of eight ethnic groups reported, Pacific islanders that represented a minority of cases (N=53; <0·1%) were pooled with Asians and those identified as “mixed” and “other” were pooled together. For descriptive analyses, countries were grouped within six world regions as defined by WHO.(20)

Information regarding cleft types in Smile Train’s records included the anatomic location (lip, alveolus, hard palate, soft palate, submucous hard/soft palate), laterality (left, right, bilateral), and completeness (complete/incomplete) of the cleft. Cases with clefts of the hard and/or soft palate also included rarer cases of submucous clefts of the palate. Isolated alveolar clefts were categorized with clefts of the palate. Cleft types, whether of the lip, alveolus, or palate, were considered regardless of their completeness. We classified clefts according to three main categories (cleft lip only [CLO], cleft palate only [CPO], and cleft lip and palate [CLP]) according to ICD-10.(21) Cases with missing information on any of the cleft anatomic location and those erroneously reported as having no clefts were excluded from analyses.

The anthropometric index weight-for-age z score was generated from weight, age, and gender variables, using WHO Stata macro.(22) The prevalence of underweight corresponded to the percentage of children with weight-for-age z scores <-2 SD away from the mean of the reference population.(23) Extreme weight-for-age z-score values were flagged as outliers according to WHO cleaning criteria, i.e. z scores either <-6 SD or >+5 SD. Patients with extreme values or missing z-scores due to missing or negative weight values were also excluded.

### Analyses

Data management and descriptive statistics were conducted using Stata/IC v14.2.(24) We seldom used small-sample statistical inference.(25) To interpret findings, we reported confidence intervals (CI) that provide a range of the magnitude of a variable of interest instead of relying on p-values. We calculated the median age at surgery, the 25^th^ percentile (Q1), and the 75th percentile (Q3). Primary surgery was considered late in children with CL±P over 1 year of age and in children with CPO over 2 years of age. For the purpose of comparison with country-specific survey data, we arbitrarily excluded countries with a total number of cases <200 (41 countries and 2,168 cases excluded) and examined the prevalence of underweight in the 60 remaining countries. The most recent estimates of underweight prevalence in children aged 0-5 years in 42 countries were extracted from the DHS program using STATcompiler.(26) For each country, a Z-test at a significance level of 5% was used to compare the prevalence of underweight in patients with cleft with the DHS estimate (assumed to be - for statistical purpose - the true population prevalence). Finally, a Spearman’s rank test was used to examine a potential correlation across countries between the prevalence of underweight in children with cleft and DHS estimates.

### Ethics

Patients or their care givers have consented to their data being captured and used by Smile Train for reviews of quality, education, evaluation, and for marketing and communication purposes. This study was approved on June 1, 2018, by the MSc Research Ethics Committee at the London School of Hygiene and Tropical Medicine (ref: 15433).

## Results

Records from 638,988 children aged ≤5 years at the time of primary cleft surgery could be retrieved from Smile Train’s database for the period between July 1, 2008 and June 30, 2018. Information on weight was missing in 4,843 cases (0·8%) and erroneously reported in 556 cases (0.03%). In consequence, there was a total of 633,589 patients for whom a weight-for-age z score could be calculated. A small percentage (3·6%) of z scores were flagged as extreme values and excluded, leaving 610,714 patients for whom z-scores could be used to calculate the prevalence of underweight (z scores <-2 SD) in the dataset. Among these patients, 602,568 had valid information on cleft types.

Females represented about 40% of all cases (**Table 1**). Nearly half (49·1%) of the children were <1 year and 76·4% were <2 years of age. About half (48·8%) of the children were identified as Asians and 36·0% as Indians; other ethnic groups in the dataset (15·2%) included Black Africans, Hispanics, Caucasians, and mixed /other groups. Children originated from 101 countries, grouped into six geographical regions (**Supplementary Table 1**). Over 75% of the children in the dataset were from South-East Asia and the Western Pacific region.

**Table 1.**
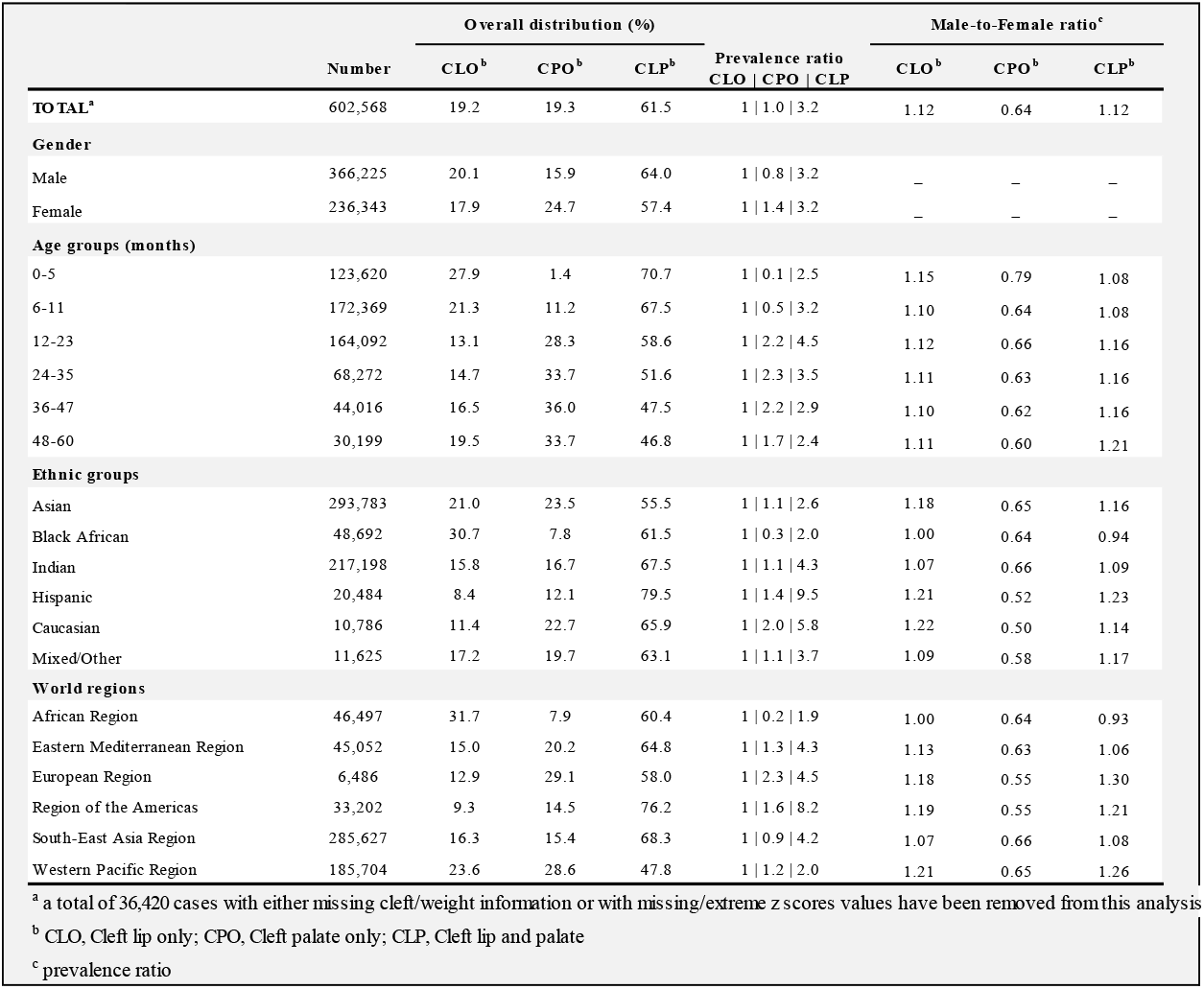
Distribution of main cleft types in records of children ≤5 years in the clinical database.

|  | Number | Overall distribution (%) |  |  | Prevalence ratio<br>CLO \| CPO \| CLP | Male-to-Female ratio<sup>c</sup> |  |  |
| --- | --- | --- | --- | --- | --- | --- | --- | --- |
|  |  | CLO<sup>b</sup> | CPO<sup>b</sup> | CLP<sup>b</sup> |  | CLO<sup>b</sup> | CPO<sup>b</sup> | CLP<sup>b</sup> |
| **TOTAL<sup>a</sup>** | 602,568 | 19.2 | 19.3 | 61.5 | 1 \| 1.0 \| 3.2 | 1.12 | 0.64 | 1.12 |
| **Gender** |  |  |  |  |  |  |  |  |
| Male | 366,225 | 20.1 | 15.9 | 64.0 | 1 \| 0.8 \| 3.2 | – | – | – |
| Female | 236,343 | 17.9 | 24.7 | 57.4 | 1 \| 1.4 \| 3.2 | – | – | – |
| **Age groups (months)** |  |  |  |  |  |  |  |  |
| 0-5 | 123,620 | 27.9 | 1.4 | 70.7 | 1 \| 0.1 \| 2.5 | 1.15 | 0.79 | 1.08 |
| 6-11 | 172,369 | 21.3 | 11.2 | 67.5 | 1 \| 0.5 \| 3.2 | 1.10 | 0.64 | 1.08 |
| 12-23 | 164,092 | 13.1 | 28.3 | 58.6 | 1 \| 2.2 \| 4.5 | 1.12 | 0.66 | 1.16 |
| 24-35 | 68,272 | 14.7 | 33.7 | 51.6 | 1 \| 2.3 \| 3.5 | 1.11 | 0.63 | 1.16 |
| 36-47 | 44,016 | 16.5 | 36.0 | 47.5 | 1 \| 2.2 \| 2.9 | 1.10 | 0.62 | 1.16 |
| 48-60 | 30,199 | 19.5 | 33.7 | 46.8 | 1 \| 1.7 \| 2.4 | 1.11 | 0.60 | 1.21 |
| **Ethnic groups** |  |  |  |  |  |  |  |  |
| Asian | 293,783 | 21.0 | 23.5 | 55.5 | 1 \| 1.1 \| 2.6 | 1.18 | 0.65 | 1.16 |
| Black African | 48,692 | 30.7 | 7.8 | 61.5 | 1 \| 0.3 \| 2.0 | 1.00 | 0.64 | 0.94 |
| Indian | 217,198 | 15.8 | 16.7 | 67.5 | 1 \| 1.1 \| 4.3 | 1.07 | 0.66 | 1.09 |
| Hispanic | 20,484 | 8.4 | 12.1 | 79.5 | 1 \| 1.4 \| 9.5 | 1.21 | 0.52 | 1.23 |
| Caucasian | 10,786 | 11.4 | 22.7 | 65.9 | 1 \| 2.0 \| 5.8 | 1.22 | 0.50 | 1.14 |
| Mixed/Other | 11,625 | 17.2 | 19.7 | 63.1 | 1 \| 1.1 \| 3.7 | 1.09 | 0.58 | 1.17 |
| **World regions** |  |  |  |  |  |  |  |  |
| African Region | 46,497 | 31.7 | 7.9 | 60.4 | 1 \| 0.2 \| 1.9 | 1.00 | 0.64 | 0.93 |
| Eastern Mediterranean Region | 45,052 | 15.0 | 20.2 | 64.8 | 1 \| 1.3 \| 4.3 | 1.13 | 0.63 | 1.06 |
| European Region | 6,486 | 12.9 | 29.1 | 58.0 | 1 \| 2.3 \| 4.5 | 1.18 | 0.55 | 1.30 |
| Region of the Americas | 33,202 | 9.3 | 14.5 | 76.2 | 1 \| 1.6 \| 8.2 | 1.19 | 0.55 | 1.21 |
| South-East Asia Region | 285,627 | 16.3 | 15.4 | 68.3 | 1 \| 0.9 \| 4.2 | 1.07 | 0.66 | 1.08 |
| Western Pacific Region | 185,704 | 23.6 | 28.6 | 47.8 | 1 \| 1.2 \| 2.0 | 1.21 | 0.65 | 1.26 |

<sup>a</sup> a total of 36,420 cases with either missing cleft/weight information or with missing/extreme z scores values have been removed from this analysis
<sup>b</sup> CLO, Cleft lip only; CPO, Cleft palate only; CLP, Cleft lip and palate
<sup>c</sup> prevalence ratio

Overall, three out of five children (61·5%) who underwent primary surgery had CLP, one in five had CLO (19·2%) and one in five had CPO (19·3%) (**Table 1**). However, the relative proportions of the three main cleft types varied with ethnicity and geography. Across all ethnic groups, over 50% of primary surgeries were in children with CLP, a phenotype at least twice as prevalent as CLO. The lowest CLP-to-CLO ratio (2:1) was found among Black African children. In contrast, the number of primary surgeries was ten times higher in Hispanic children with CLP than in those with CLO. Further, only 7·9% of the primary surgeries in Africa were conducted in children with CPO (largely identified as Black Africans) whereas this group represented about 15% of all primary cleft surgeries in Latin America (primarily in Hispanics and Caucasians) and South-East Asia (primarily in Indians and Asians), 20% in the Eastern Mediterranean region (primarily in Asians and Indians), and close to 30% in the Western Pacific region (largely in Asians) and European region (primarily in Caucasians and Asians). We also observed gender differences in cleft types. Children with CPO were more likely to be females (sex prevalence ratio of 0·6) whereas there was a slight predominance of males among children with either CLO or CLP (sex prevalence ratio of 1·2). This pattern was true across age groups, ethnic groups, and world regions (**Table 1**).

The median age at the time of primary surgery was greater in children with CPO (∼20 months) compared to those with CLO (∼8·5 months) or CLP (∼10·5 months) (**Table 2**). When considering primary surgeries completed in children aged over 1 year, there were no gender differences, yet some variations across ethnic groups and world regions (**Table 2**). For instance, a greater percentage of children with CLO were operated after the age of 1 year in Africa (47·9%) and South-East Asia (45·9%) compared to other settings. Depending on the setting, 40·7% to 46·6% of children with CLP underwent primary surgery after 1 year of age and 33·6% to 47·2% of children with CPO after 2 years of age.

**Table 2.** Median age at the time of surgery and prevalence of late primary operations across cleft types, gender, ethnicities, and world regions.

|  | Median age at surgery (Q<sub>1</sub>,Q<sub>3</sub>) in years |  |  | % Late primary surgeries (95% CI) |  |  |
| --- | --- | --- | --- | --- | --- | --- |
|  | CLO<sup>a</sup> | CPO<sup>a</sup> | CLP<sup>a</sup> | CLO (>1 year) | CPO (>2 years) | CLP (>1 year) |
| **TOTAL<sup>a</sup>** | 0.72 (0.45,1.55) | 1.68 (1.10,2.82) | 0.88 (0.51,1.60) | 37.9 (37.6-38.1) | 41.6 (41.3-41.9) | 44.3 (44.1-44.4) |
| **Gender** |  |  |  |  |  |  |
| Male | 0.71 (0.44,1.55) | 1.66 (1.10,2.78) | 0.90 (0.51,1.62) | 37.8 (37.4-38.1) | 40.9 (40.5-41.3) | 44.9 (44.7-45.1) |
| Female | 0.72 (0.45,1.55) | 1.71 (1.10,2.86) | 0.85 (0.50,1.56) | 38.0 (37.6-38.5) | 42.4 (42.0-42.8) | 43.1 (42.8-43.4) |
| **Ethnic groups** |  |  |  |  |  |  |
| Asian | 0.64 (0.42,1.29) | 1.84 (1.21,2.96) | 0.92 (0.50,1.71) | 32.7 (32.3-33.1) | 45.4 (45.1-45.8) | 46.2 (46.0-46.5) |
| Black African | 0.94 (0.47,2.20) | 1.70 (1.03,2.87) | 0.77 (0.42,1.64) | 47.7 (46.9-46.5) | 42.2 (40.7-43.8) | 41.0 (40.4-41.5) |
| Indian | 0.86 (0.51,1.98) | 1.48 (1.01,2.56) | 0.88 (0.53,1.52) | 44.5 (44.0-45.0) | 36.3 (35.8-36.8) | 43.5 (43.2-43.7) |
| Hispanic | 0.53 (0.36,0.97) | 1.61 (1.10,2.45) | 0.82 (0.43,1.48) | 24.1 (22.1-26.1) | 36.3 (34.4-38.3) | 41.7 (41.0-42.5) |
| Caucasian | 0.50 (0.35,0.82) | 1.38 (1.03,2.15) | 0.77 (0.43,1.36) | 20.1 (17.9-22.4) | 27.9 (26.1-29.7) | 41.9 (40.8-43.1) |
| Mixed/Other | 0.59 (0.35,1.35) | 1.42 (0.97,2.35) | 0.72 (0.39,1.36) | 32.6 (30.6-34.7) | 30.0 (28.1-31.9) | 36.9 (35.8-38.0) |
| **World regions** |  |  |  |  |  |  |
| African Region | 0.93 (0.47,2.18) | 1.69 (1.03,2.84) | 0.76 (0.41,1.62) | 47.4 (46.6-48.2) | 42.1 (40.5-43.7) | 40.2 (39.6-40.8) |
| Eastern Mediterranean Region | 0.74 (0.42,2.00) | 1.49 (1.00,2.58) | 0.84 (0.48,1.56) | 40.3 (39.1-41.4) | 36.5 (35.5-37.5) | 41.8 (41.3-42.4) |
| European Region | 0.58 (0.41,1.01) | 1.69 (1.12,2.97) | 0.85 (0.51,1.49) | 25.4 (22.5-28.4) | 40.5 (38.3-42.7) | 42.9 (41.3-44.5) |
| Region of the Americas | 0.53 (0.36,0.96) | 1.56 (1.11,2.37) | 0.82 (0.43,1.49) | 23.6 (22.1-25.1) | 33.3 (31.9-34.6) | 42.6 (42.0-43.2) |
| South-East Asia Region | 0.88 (0.52,1.99) | 1.51 (1.02,2.61) | 0.90 (0.53,1.57) | 45.2 (44.8-45.7) | 37.3 (36.8-37.8) | 44.6 (44.4-44.8) |
| Western Pacific Region | 0.59 (0.40,1.09) | 1.89 (1.26,2.98) | 0.90 (0.49,1.72) | 27.7 (27.3-28.1) | 46.8 (46.4-47.2) | 46.1 (45.8-46.5) |
| <sup>a</sup> 602,568 cases |  |  |  |  |  |  |
| <sup>b</sup> CLO, Cleft lip only; CPO, Cleft palate only; CLP, Cleft lip and palate |  |  |  |  |  |  |

Overall, 28·6% of children had a low weight-for-age z score (<-2 SD) at the time of surgery (**Table 3**). The prevalence of underweight varied with gender, age at surgery, ethnicity/geography, and type of cleft. Underweight was more prevalent in children with CLP (32·8% vs. 25·5% with CLO and 18·4% with CPO), in male cases (30·9% vs. 25·0% in females), in older children (33·6% at 48-60 months vs. 26·9% at 0-5 months), and in children from South-East Asia (40·4%) – primarily Indians (43·7%). We noticed that the prevalence of underweight at surgery was systematically lower in children with CPO compared to those with CL±P, regardless of gender, age at surgery, and ethnicity – except among Black Africans (**Table 3**).

**Table 3.** Prevalence of underweight at surgery according to the type of cleft and across gender, age groups, ethnicities, and world regions.

|  | Weight-for-age z score <-2 SD<br>% (95% CI)<sup>b</sup> |  |  |  |
| --- | --- | --- | --- | --- |
|  | CLO<sup>c</sup> | CPO<sup>c</sup> | CLP<sup>c</sup> | Any clefts |
| **TOTAL<sup>a</sup>** | 25.5 (25.2-25.7) | 18.4 (18.2-18.6) | 32.8 (32.6-32.9) | 28.6 (28.5-28.7) |
| **Gender** |  |  |  |  |
| Male | 26.9 (26.6-27.2) | 20.0 (19.7-20.4) | 34.8 (34.6-35.0) | 30.9 (30.7-31.0) |
| Female | 23.0 (22.6-23.4) | 16.8 (16.5-17.1) | 29.2 (29.0-29.5) | 25.0 (24.9-25.2) |
| **Age groups** |  |  |  |  |
| 0-5 mo | 17.0 (16.6-17.4) | 14.2 (12.6-15.9) | 31.1 (30.8-31.4) | 26.9 (26.7-27.2) |
| 6-11 mo | 23.6 (23.2-24.1) | 16.8 (16.3-17.3) | 34.9 (34.6-35.1) | 30.5 (30.2-30.7) |
| 12-23 mo | 29.6 (29.0-30.2) | 14.3 (14.0-14.6) | 28.6 (28.3-28.9) | 24.7 (24.5-24.9) |
| 24-35 mo | 37.0 (36.1-37.9) | 20.6 (20.0-21.1) | 35.7 (35.2-36.2) | 30.8 (30.4-31.1) |
| 36-47 mo | 38.7 (37.6-39.9) | 24.5 (23.8-25.2) | 39.1 (38.5-39.8) | 33.8 (33.4-34.2) |
| 48-60 mo | 36.2 (34.9-37.4) | 26.4 (25.5-27.3) | 37.8 (37.0-38.6) | 33.6 (33.1-34.2) |
| **Ethnic group** |  |  |  |  |
| Asian | 15.8 (15.5-16.1) | 11.4 (11.1-11.6) | 23.0 (22.8-23.2) | 18.7 (18.6-18.9) |
| Black African | 24.3 (23.6-25.0) | 24.8 (23.4-26.2) | 32.6 (32.1-33.2) | 29.5 (29.1-29.9) |
| Indian | 44.3 (43.8-44.8) | 32.5 (32.0-33.0) | 46.3 (46.0-46.6) | 43.7 (43.5-43.9) |
| Hispanic | 17.4 (15.7-19.3) | 13.3 (12.0-14.7) | 21.5 (20.9-22.1) | 20.2 (19.6-20.7) |
| Caucasian | 17.3 (15.3-19.5) | 10.0 (8.9-11.3) | 15.4 (14.6-16.3) | 14.4 (13.8-15.1) |
| Mixed/Other | 22.7 (20.9-24.6) | 12.2 (11.0-13.6) | 22.6 (21.6-23.5) | 20.6 (19.8-21.3) |
| **World region of origin** |  |  |  |  |
| African region | 24.3 (23.6-25.0) | 25.1 (23.7-26.5) | 32.6 (32.0-33.1) | 29.4 (28.9-29.8) |
| Eastern Mediterranean region | 32.1 (31.0-33.2) | 21.9 (21.1-22.8) | 39.3 (15.0-17.5) | 34.7 (34.2-35.1) |
| European region | 11.9 (9.9-14.3) | 8.0 (6.9-9.3) | 12.4 (11.3-13.4) | 11.0 (10.3-11.8) |
| Region of the Americas | 16.2 (15.0-17.5) | 11.9 (11.0-12.8) | 18.5 (18.0-19.0) | 17.3 (16.9-17.7) |
| South-East Asia region | 39.9 (39.5-40.4) | 31.6 (31.2-32.1) | 42.5 (42.3-42.8) | 40.4 (40.3-40.6) |
| Western Pacific region | 10.4 (10.1-10.7) | 7.4 (7.2-7.6) | 14.2 (13.9-14.4) | 11.3 (11.2-11.5) |
| <sup>a</sup> 602,568 cases |  |  |  |  |
| <sup>b</sup> percentage of underweight children |  |  |  |  |
| <sup>c</sup> CLO, Cleft lip only; CPO, Cleft palate only; CLP, Cleft lip and palate |  |  |  |  |

**Table 4** displays the prevalence of underweight across the main cleft types in children from 60 LMICs. Mean weight-for-age z scores ± SD are available as supplementary information (**Supplementary Table 2**). The prevalence of underweight among children at the time of surgery was either high (between 20-29%) or very high (≥30%) in two thirds of all 60 countries. Single-point estimates for underweight prevalence in children ≤5 years from DHS could be retrieved for all 21 African countries, yet only for 4 out of 7 countries in the Eastern Mediterranean region, 3 out of 4 in the European region, 7 out of 12 in Latin America, 6 out of 7 in South-East Asia, and only 1 out of 6 in the Western Pacific region (**Table 4**). Using DHS values as population estimates for the prevalence of underweight in children without cleft, we compared the prevalence of underweight in cleft patients to DHS estimates in 42 countries. The prevalence of underweight was higher in children with cleft than in those without cleft (p<0·05) except in Burundi, Ethiopia, Yemen, and Uzbekistan. In these countries, the DHS estimates for underweight prevalence in the population were above the prevalence of underweight in children with cleft (p≤0·0001). In addition, there was no evidence of a difference in underweight prevalence between children with cleft and the DHS estimate in Niger (p=0·74). Finally, we found evidence of a correlation between the prevalence of underweight in the DHS program and the prevalence of underweight among children with cleft (**Figure 1**; any clefts, r_s_: 0·6305; p<0·0001). The positive correlation persisted when the analysis was restricted to either children with CLO (r_s_: 0·5945, p<0·0001), CPO (r_s_: 0·5157, p<0·001), or CLP (r_s_: 0·5634, p=0·0001).

**Table 4.**
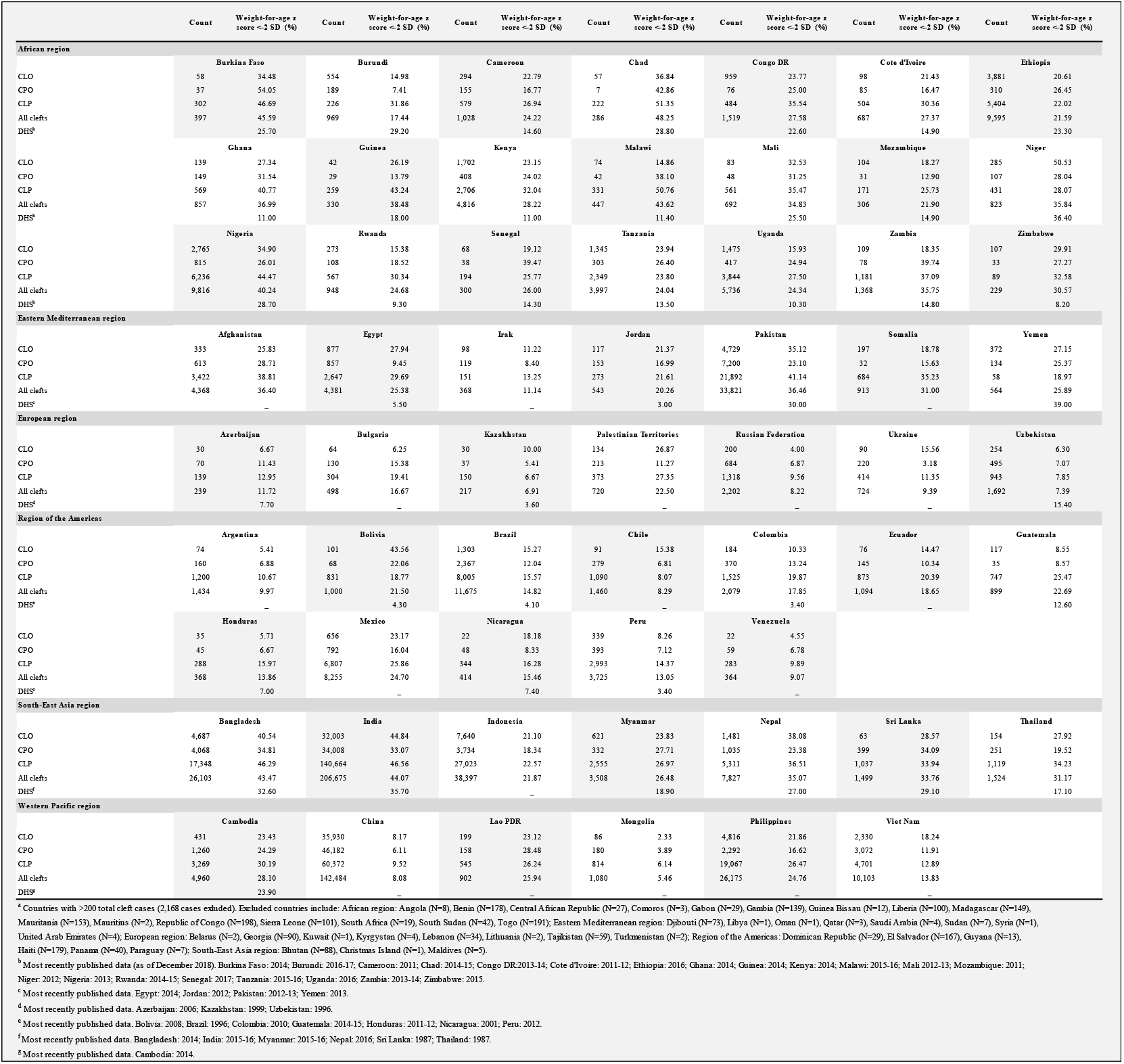
Prevalence of underweight at the time of primary cleft surgery in 60 countries^a^.

**Fig 1.**
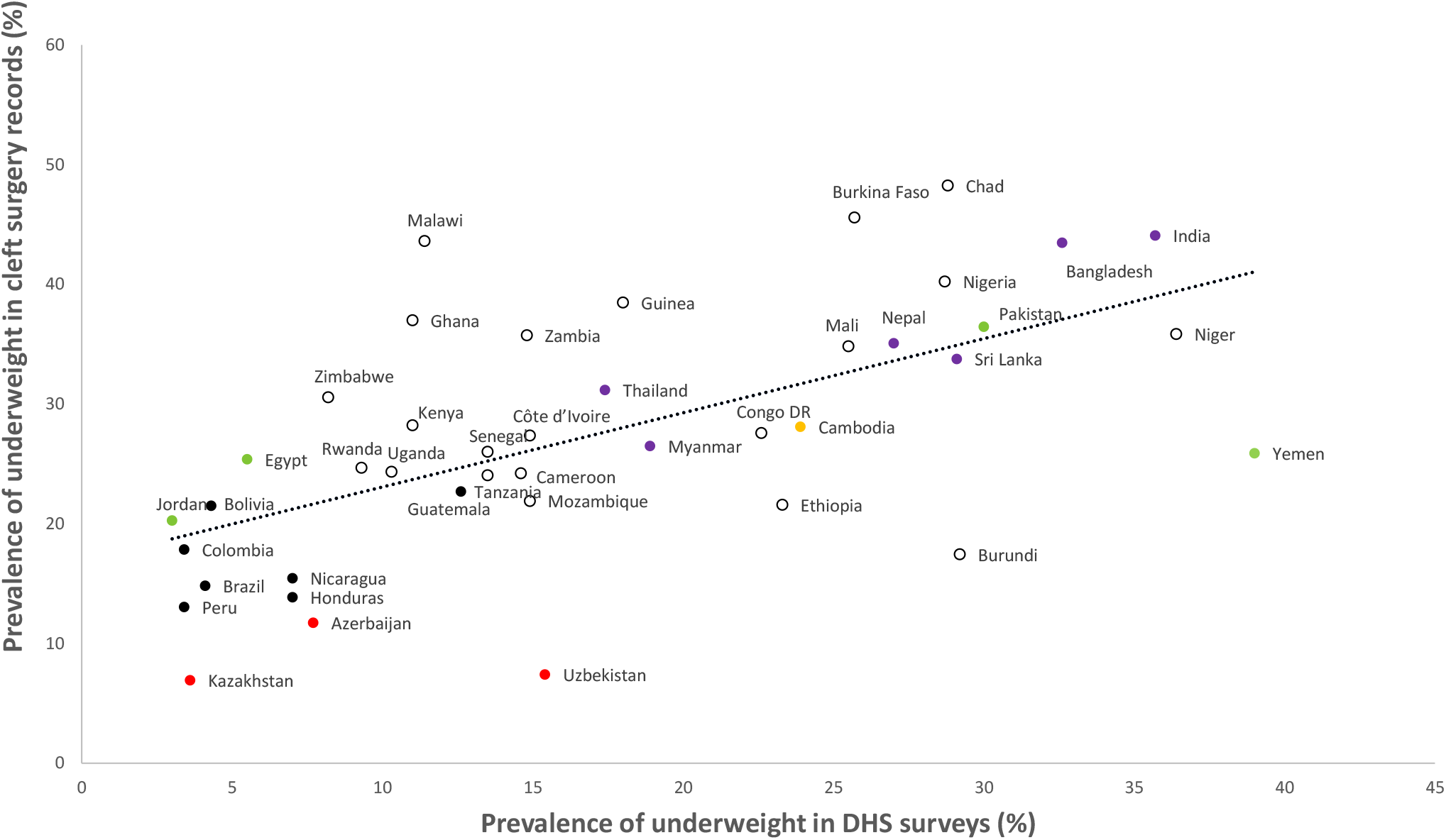
Correlation of underweight prevalences in clinical records and national surveys across 42 countries.

## Discussion

The analysis of over 600,000 records of primary cleft operations conducted in LMICs over a 10 year-period showed that 28·6% of children were underweight at the time of primary surgery. This figure is well above the global underweight prevalence in children ≤5 years estimated at 13·5% in 2017(27) and may reflect the additional unmet needs of children with cleft in settings of widespread malnutrition.

Orofacial cleft is not considered to be a life-threatening condition requiring emergency surgery. However, our findings suggest that children born with a cleft in limited-resource settings are at high risk of failure to thrive. The limited evidence of malnutrition in children with unrepaired cleft in LMICs has come so far from small observational studies with significant heterogeneity especially in anthropometry methods of assessing nutritional status (i.e. use of different reference growth curves, different anthropometric indices, and different cut-offs to estimate the prevalence of malnutrition).(7-9, 11-13, 15) Out of the two studies that used weight-for-age z score with a cut-off of -2 SD but reportedly used the NCHS reference growth curves(7, 15), one conducted in Nigeria compared the nutritional status of 50 children with cleft and 50 peers without cleft.(7) The authors reported that 26% of the children with cleft were underweight, yet found no evidence of a difference with children without cleft. The other study led by Lazarus and colleagues (15) two decades ago in South Africa reported high prevalence estimates of underweight in 640 children with cleft at the time of primary surgery: 21·8% with CLO, 36·1% with CPO, and 29·6% with (unilateral) CLP.

Failure to thrive from birth is sometimes explained in children with isolated and syndromic CP±L, Pierre Robin Sequence, and bilateral/complete CL±P by their prevailing feeding difficulties.(6) In contrast, unilateral CLO is usually thought to have little impact on the feeding process.(3, 8) Yet, our findings suggest that children with CLO were not less likely than those with CP±L to be underweight at the time of surgery, even among those operated within the first 6 months of life. Besides, regardless of the type of cleft, comparisons with DHS estimates across countries showed that children with cleft were more likely to be underweight, and thus faced more challenges to thrive, than their peers without cleft. There is a paucity of data on the impact of cleft on the care and safety of children in LMICs. Some evidence highlights stigma due to lack of awareness and misconceptions as a major barrier to care for children with cleft.(28) Child neglect may be especially affecting those with CL±P as the cleft of the lip is prominent and disfiguring. Additional challenges known to prevent timely and appropriate care of neonates and infants with disability in low-resource settings can also explain a higher prevalence of malnutrition among children with unrepaired cleft.(29, 30) Barriers related to sociocultural norms, lack of knowledge and awareness, distance to health facilities, cost of healthcare, lack of specialist feeding support, family structure, household income, and maternal factors (education, health, responsibilities, etc.) may be disproportionally affecting children with cleft compared to their peers without cleft. Immediate specialist attention from birth would thus be paramount to overcome many of these barriers and reduce the risk of failure to thrive in these children.

Disparities in the nutrition situation of populations across LMICs may account for the observed differences in the prevalence of underweight in children with cleft across countries.(31) Globally, we found a higher prevalence of underweight in children with cleft in countries with higher prevalence estimates of underweight children in the general population. Besides, inadequate infrastructures and difficulties accessing care, including surgery, may also be variously contributing to the risk of malnutrition and to delays in cleft repair. For each cleft type and across settings, we found however relatively limited differences in the median age at primary surgery and, accordingly, limited variations in the percentages of late primary surgeries. In contrast, there were substantial differences in the relative proportions of cleft types across ethnic groups and settings. These differences might explain some of the variations in the prevalence of underweight at surgery. Indeed, with children with complex/severe cases (i.e. CLP cases) being more likely to be underweight, a higher proportion of CLP-to-CLO cases contributes to a higher overall prevalence of malnutrition in a given setting.(3, 12, 15) We also reported large variations in the frequency of CPO across settings and ethnic groups, with a frequency three times higher among Caucasians and Asians than among Black Africans. Studies have previously reported a low frequency of CPO in Sub-Saharan Africa.(32-36) A cleft of the palate affects the normal sucking process, placing children with CP±L at immediate risk of undernutrition and failure to thrive.(3, 37) This is especially true in the absence of timely specialist feeding assistance.(38) However, overall as well as across most ethnic groups and settings, children with CPO at surgery were less likely to be underweight than those with CL±P. Several reasons could explain these observations. The timing of lip and palate repairs differs such that lips of children with CL±P are repaired from the age of 3 months whereas surgeons are unlikely to repair a palate before 6 months of age, and some may wait until children are 12 or 18 months of age. The waiting time may allow for feeding and nutrition counselling to correct or prevent undernutrition. Yet, in many settings, children with cleft are unlikely to present at hospitals unless they have been actively searched for and identified in the community. CPO can be easily missed, especially in settings with little-to-no postnatal check-up and/or no awareness of cleft.(36, 39) Moreover, besides the struggle to feed, children with CPO are more likely than those with CL±P to have associated congenital anomalies that may further threaten their survival.(40) (41) Thus, we cannot exclude that children with CPO being operated in many settings are among the mildest cases, that is more likely to have survived the lack of specialist care and less likely to be undernourished.(36) Finally, we also reported a lower prevalence of underweight among females across all age groups, ethnic groups, and settings, suggesting that biological differences rather than gender inequality in accessing nutrition and health care might play a role.(9) This difference between gender may also contribute to differences in the prevalence of underweight between cleft types, with CPO affecting about 1.5 times more females than males.(42)

This study is the first to examine the scale of the malnutrition burden in children with cleft across lower-resource countries where immediate care at birth and access to cleft surgery are limited. Yet, our overall and country-specific findings only account for children who have been operated by Smile Train’s partners. Children who have not been identified, those who have been lost to follow-up before being operated, and those operated by other surgical teams are not accounted for. However, although the records are limited to clinical data, the relative proportions of cleft types in this database are reasonably consistent with the current understanding of cleft epidemiology across ethnic groups and settings.(42, 43) Figures may also underestimate the scale of malnutrition because a substantial proportion of the children operated by Smile Train’s partners may have received some level of feeding and nutritional care prior to surgery to promote weight gain, correct anaemia, and ensure fitness to surgery. Another limitation of the present study relates to the use of weight-for-age z scores to estimate the nutritional status of children with cleft. Weight-for-age is a composite index that cannot distinguish between different forms of malnutrition,(44) and additional anthropometric indicators are needed to establish the prevalence of stunting and/or wasting.(45) Another limitation with the use of weight-for-age z scores is that the data based on this index may be biased by inaccuracies in dates of birth and weight measurements (44). Finally, the use of country-specific DHS estimates as an approximation of the nutritional situation of (non-cleft) children ≤5 years old may be limited by the fact that (i) survey data are captured at specific points in time, whereas we estimated the prevalence of underweight using clinical data collected over a 10 year-period, (ii) the age distribution in our dataset may not match the age distribution in DHS survey samples, and (iii) national estimates were not available for all countries and in some cases (e.g. Sri Lanka) were not up-to-date. Nonetheless, DHS data provided reliable country-specific estimates of underweight prevalence readily available for comparisons.

Our findings highlight the urgent need of initiatives on child and maternal health that will equip parents, communities, birth attendants, midwifes, nutritionists, and other healthcare professionals in LMICs with the necessary knowledge and skills that will ensure prompt feeding care and specialist attention for children with cleft. Poverty, low awareness and /education, social neglect, lack of immediate feeding support and unhealthy feeding practices, lack of access to pediatric and surgical care, and absence of birth defect registries are among the many issues that need to be addressed to give children with cleft in LMICs the chance to survive and develop to their full potential. Efforts to protect the most disadvantaged children, including those born with a cleft, are necessary to achieve significant progress towards the health targets of SDG 3(46) and concomitantly reduce health inequalities within countries.

## Data Availability

Access to the original dataset needs to be requested directly to cleft NGO Smile Train.

## Abbreviations

CLO: Cleft lip only;
CLP: Cleft lip and palate;
CL±P: Cleft lip with or without cleft palate;
CP±L: Cleft palate with or without cleft lip;
CPO: Cleft palate only;
DHS: Demographic and Health Surveys;
ICD-10: International Classification of Diseases, 10^th^ edition;
LMICs: Low- and middle-income countries;
NCHS: National Center for Health Statistics;
WHO: World Health Organization.

## Contributions

BD, MK, and PS conceived the research project. PS extracted the underlying data of interest from Smile Train digital patient database. Both BD and MK have verified the underlying data and contributed to the analysis design. BD executed the analyses, interpreted the findings, wrote the first draft, revised subsequent draft, and prepare the manuscript for submission. MK, PS, and ES revised drafts.

## Acknowledgments

We are grateful to all Smile Train cleft partners worldwide for collecting the data included in this study. We also want to thank all Smile Train partners in Africa who contributed to improve data interpretation by sharing their expertise.

---

**S1 Table.**
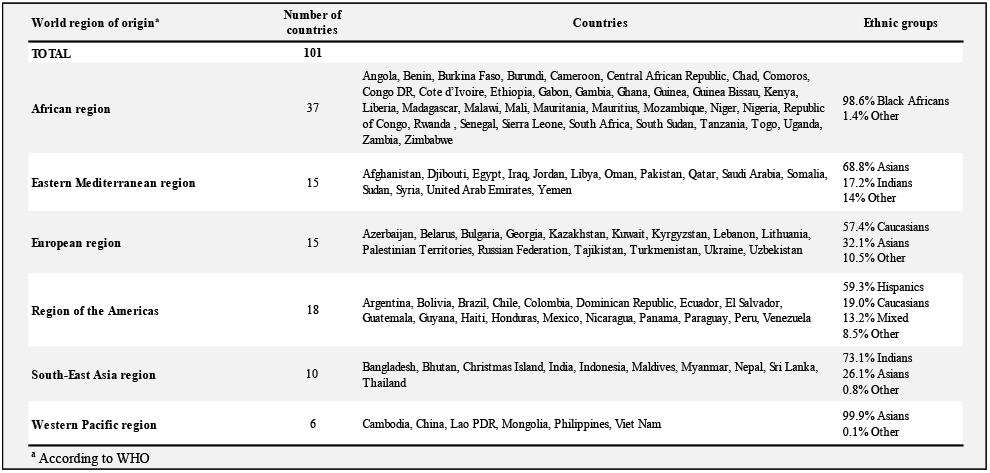
Geographic distribution and ethnicity of cleft cases (N=602,568).

| World region of origin<sup>a</sup> | Number of countries | Countries | Ethnic groups |
| --- | --- | --- | --- |
| TOTAL | 101 |  |  |
| African region | 37 | Angola, Benin, Burkina Faso, Burundi, Cameroon, Central African Republic, Chad, Comoros, Congo DR, Cote d'Ivoire, Ethiopia, Gabon, Gambia, Ghana, Guinea, Guinea Bissau, Kenya, Liberia, Madagascar, Malawi, Mali, Mauritania, Mauritius, Mozambique, Niger, Nigeria, Republic of Congo, Rwanda, Senegal, Sierra Leone, South Africa, South Sudan, Tanzania, Togo, Uganda, Zambia, Zimbabwe | 98.6% Black Africans<br>1.4% Other |
| Eastern Mediterranean region | 15 | Afghanistan, Djibouti, Egypt, Iraq, Jordan, Libya, Oman, Pakistan, Qatar, Saudi Arabia, Somalia, Sudan, Syria, United Arab Emirates, Yemen | 68.8% Asians<br>17.2% Indians<br>14% Other |
| European region | 15 | Azerbaijan, Belarus, Bulgaria, Georgia, Kazakhstan, Kuwait, Kyrgyzstan, Lebanon, Lithuania, Palestinian Territories, Russian Federation, Tajikistan, Turkmenistan, Ukraine, Uzbekistan | 57.4% Caucasians<br>32.1% Asians<br>10.5% Other |
| Region of the Americas | 18 | Argentina, Bolivia, Brazil, Chile, Colombia, Dominican Republic, Ecuador, El Salvador, Guatemala, Guyana, Haiti, Honduras, Mexico, Nicaragua, Panama, Paraguay, Peru, Venezuela | 59.3% Hispanics<br>19.0% Caucasians<br>13.2% Mixed<br>8.5% Other |
| South-East Asia region | 10 | Bangladesh, Bhutan, Christmas Island, India, Indonesia, Maldives, Myanmar, Nepal, Sri Lanka, Thailand | 73.1% Indians<br>26.1% Asians<br>0.8% Other |
| Western Pacific region | 6 | Cambodia, China, Lao PDR, Mongolia, Philippines, Viet Nam | 99.9% Asians<br>0.1% Other |
| <sup>a</sup> According to WHO |  |  |  |

**S2 Table.** Mean weight-for-age z scores and prevalences of underweight at primary cleft surgery in 60 countries^a^.

|  | Weight-for-age Z scores |  |  | Weight-for-age Z scores |  |  | Weight-for-age Z scores |  |  | Weight-for-age Z scores |  |  | Weight-for-age Z scores |  |  | Weight-for-age Z scores |  |  | Weight-for-age Z scores |  |  |
| --- | --- | --- | --- | --- | --- | --- | --- | --- | --- | --- | --- | --- | --- | --- | --- | --- | --- | --- | --- | --- | --- |
|  | Count (%) | Mean (SD) | <2 SD z score | Count (%) | Mean (SD) | % | Count (%) | Mean (SD) | % | Count (%) | Mean (SD) | % | Count (%) | Mean (SD) | % | Count (%) | Mean (SD) | % | Count (%) | Mean (SD) | % |
| African region |  |  |  |  |  |  |  |  |  |  |  |  |  |  |  |  |  |  |  |  |  |
|  | Burkina Faso |  |  | Burundi |  |  | Cameroon |  |  | Chad |  |  | Congo DR |  |  | Cote d'Ivoire |  |  | Ethiopia |  |  |
| Cleft lip only | 58 (15) | -1.36 (1.48) | 34.48 | 554 (57) | 0.10 (2.00) | 14.98 | 294 (29) | -0.55 (1.99) | 22.79 | 57 (20) | -1.29 (1.55) | 36.84 | 959 (63) | -0.78 (1.79) | 23.77 | 98 (14) | -0.99 (1.58) | 21.43 | 3,881 (40) | -0.84 (1.51) | 20.61 |
| Cleft palate only | 37 (9) | -2.17 (1.65) | 54.05 | 189 (20) | 1.03 (2.13) | 7.41 | 155 (15) | -0.15 (1.91) | 16.77 | 7 (2) | -1.37 (2.41) | 42.86 | 76 (5) | -0.86 (1.87) | 25.00 | 85 (12) | -0.49 (1.78) | 16.47 | 310 (3) | -0.87 (1.57) | 26.45 |
| Cleft lip and palate | 302 (76) | -1.95 (1.59) | 46.69 | 226 (23) | -0.87 (2.09) | 31.86 | 579 (56) | -0.88 (1.94) | 26.94 | 222 (78) | -2.02 (1.78) | 51.35 | 484 (32) | -1.27 (1.85) | 35.54 | 504 (73) | -1.20 (1.62) | 30.36 | 5,404 (56) | -0.84 (1.59) | 22.02 |
| All clefts | 397 (100) | -1.89 (1.59) | 45.59 | 969 (100) | 0.05 (2.14) | 17.44 | 1,028 (100) | -0.68 (1.96) | 24.22 | 286 (100) | -1.86 (1.77) | 48.25 | 1,519 (100) | -0.94 (1.26) | 27.58 | 687 (100) | -1.08 (1.65) | 27.37 | 9,595 (100) | -0.84 (1.56) | 21.59 |
|  | Ghana |  |  | Guinea |  |  | Kenya |  |  | Malawi |  |  | Mali |  |  | Mozambique |  |  | Niger |  |  |
| Cleft lip only | 139 (16) | -0.88 (1.86) | 27.34 | 42 (13) | -1.00 (1.48) | 26.19 | 1,702 (35) | -0.86 (1.65) | 23.15 | 74 (17) | -0.53 (1.55) | 14.86 | 83 (12) | -1.05 (1.69) | 32.53 | 104 (34) | -0.22 (1.98) | 18.27 | 285 (35) | -1.95 (1.72) | 50.53 |
| Cleft palate only | 149 (17) | -1.12 (1.91) | 31.54 | 29 (9) | -0.73 (1.54) | 13.79 | 408 (8) | -0.88 (1.70) | 24.02 | 42 (9) | -1.22 (1.59) | 38.10 | 48 (7) | -0.85 (1.77) | 31.25 | 31 (10) | -0.04 (1.60) | 12.90 | 107 (13) | -1.09 (1.74) | 28.04 |
| Cleft lip and palate | 569 (66) | -1.42 (1.88) | 40.77 | 259 (78) | -1.72 (1.61) | 43.24 | 2,706 (56) | -1.26 (1.79) | 32.04 | 331 (74) | -2.04 (1.94) | 50.76 | 561 (81) | -1.29 (1.75) | 35.47 | 171 (56) | -0.88 (1.72) | 25.73 | 431 (52) | -0.11 (1.60) | 28.07 |
| All clefts | 857 (100) | -1.28 (1.89) | 36.99 | 330 (100) | -1.54 (1.62) | 38.48 | 4,816 (100) | -1.08 (1.74) | 28.22 | 447 (100) | -1.72 (1.94) | 43.62 | 692 (100) | -1.23 (1.75) | 34.83 | 306 (100) | -0.57 (1.83) | 21.90 | 823 (100) | -1.40 (1.71) | 35.84 |
|  | Nigeria |  |  | Rwanda |  |  | Senegal |  |  | Tanzania |  |  | Uganda |  |  | Zambia |  |  | Zimbabwe |  |  |
| Cleft lip only | 2765 (28) | -1.18 (1.97) | 34.90 | 273 (29) | -0.73 (1.49) | 15.38 | 68 (23) | -0.50 (1.64) | 19.12 | 1,345 (34) | -0.52 (2.11) | 23.94 | 1,475 (26) | 0.09 (2.22) | 15.93 | 109 (8) | -0.74 (1.76) | 18.35 | 107 (47) | -1.34 (1.80) | 29.91 |
| Cleft palate only | 815 (8) | -0.75 (2.01) | 26.01 | 108 (11) | -0.69 (1.76) | 18.52 | 38 (13) | -0.98 (2.015) | 39.47 | 303 (8) | -0.84 (1.93) | 26.40 | 417 (7) | -0.25 (2.48) | 24.94 | 78 (6) | -1.59 (1.90) | 39.74 | 33 (14) | -0.77 (1.71) | 27.27 |
| Cleft lip and palate | 6,236 (64) | -1.65 (1.90) | 44.47 | 567 (60) | -1.17 (1.69) | 30.34 | 194 (65) | -0.86 (1.95) | 25.77 | 2,349 (59) | -0.33 (2.32) | 23.80 | 3,844 (67) | -0.34 (2.57) | 27.50 | 1,181 (86) | -1.56 (1.48) | 37.09 | 89 (39) | -1.31 (1.67) | 32.58 |
| All clefts | 9,816 (100) | -1.45 (1.95) | 40.24 | 948 (100) | -0.99 (1.65) | 24.68 | 300 (100) | -0.79 (1.91) | 26.00 | 3,997 (100) | -0.43 (2.22) | 24.04 | 5,736 (100) | -0.22 (2.49) | 24.34 | 1,368 (100) | -1.50 (1.55) | 35.75 | 229 (100) | -1.24 (1.74) | 30.57 |
| Eastern Mediterranean region |  |  |  |  |  |  |  |  |  |  |  |  |  |  |  |  |  |  |  |  |  |
|  | Afghanistan |  |  | Egypt |  |  | Iraq |  |  | Jordan |  |  | Pakistan |  |  | Somalia |  |  | Yemen |  |  |
| Cleft lip only | 333 (8) | -1.15 (1.50) | 25.83 | 877 (20) | -1.05 (1.67) | 27.94 | 98 (27) | 0.23 (2.01) | 11.22 | 117 (22) | -0.74 (1.51) | 21.37 | 4,729 (14) | -1.20 (1.90) | 35.12 | 197 (22) | -0.56 (1.68) | 18.78 | 372 (66) | -1.02 (1.64) | 27.15 |
| Cleft palate only | 613 (14) | -1.24 (1.53) | 28.71 | 857 (20) | -0.27 (1.41) | 9.45 | 119 (32) | 0.04 (1.57) | 8.40 | 153 (28) | -0.33 (1.84) | 16.99 | 7,200 (21) | -0.75 (1.82) | 23.10 | 32 (4) | -0.92 (1.29) | 15.63 | 134 (24) | -1.03 (1.48) | 25.37 |
| Cleft lip and palate | 3,422 (78) | -1.55 (1.64) | 38.81 | 2,647 (60) | -1.23 (1.70) | 29.69 | 151 (41) | 0.41 (2.33) | 13.25 | 273 (50) | -0.72 (1.61) | 21.61 | 21,892 (65) | -1.44 (1.96) | 41.14 | 684 (75) | -1.39 (1.95) | 35.23 | 58 (10) | -0.66 (1.79) | 18.97 |
| All clefts | 4,368 (100) | -1.48 (1.62) | 36.40 | 4,381 (100) | -1.01 (1.68) | 25.38 | 368 (100) | 0.24 (2.03) | 11.14 | 543 (100) | -0.62 (1.67) | 20.26 | 33,821 (100) | -1.26 (1.94) | 36.46 | 913 (100) | -1.20 (1.91) | 31.00 | 564 (100) | -0.98 (1.62) | 25.89 |
| European region |  |  |  |  |  |  |  |  |  |  |  |  |  |  |  |  |  |  |  |  |  |
|  | Azerbaijan |  |  | Bulgaria |  |  | Kazakhstan |  |  | Palestinian Territories |  |  | Russian Federation |  |  | Ukraine |  |  | Uzbekistan |  |  |
| Cleft lip only | 30 (13) | -0.59 (1.32) | 6.67 | 64 (13) | -0.62 (0.99) | 6.25 | 30 (14) | 0.70 (2.02) | 10.00 | 134 (19) | -0.82 (1.86) | 26.87 | 200 (9) | 0.05 (1.44) | 4.00 | 90 (12) | -0.50 (1.77) | 15.56 | 254 (15) | 0.17 (1.41) | 6.30 |
| Cleft palate only | 70 (29) | -0.15 (1.62) | 11.43 | 130 (26) | -0.66 (1.38) | 15.38 | 37 (17) | 0.35 (1.94) | 5.41 | 213 (30) | -0.11 (1.61) | 11.27 | 684 (31) | -0.06 (1.41) | 6.87 | 220 (30) | 0.53 (1.27) | 3.18 | 495 (29) | 0.15 (1.53) | 7.07 |
| Cleft lip and palate | 139 (58) | -0.58 (1.34) | 12.95 | 304 (61) | -0.96 (1.38) | 19.41 | 150 (69) | -0.20 (1.25) | 6.67 | 373 (52) | -0.87 (1.81) | 27.35 | 1,318 (60) | -0.31 (1.42) | 9.56 | 414 (57) | -0.16 (1.60) | 11.35 | 943 (56) | -0.03 (1.54) | 7.85 |
| All clefts | 239 (100) | -0.45 (1.43) | 11.72 | 498 (100) | -0.83 (1.34) | 16.67 | 217 (100) | 0.02 (1.54) | 6.91 | 720 (100) | -0.63 (1.79) | 22.50 | 2,202 (100) | -0.20 (1.43) | 8.22 | 724 (100) | 0.01 (1.57) | 9.39 | 1,692 (100) | 0.05 (1.52) | 7.39 |
| Region of the Americas |  |  |  |  |  |  |  |  |  |  |  |  |  |  |  |  |  |  |  |  |  |
|  | Argentina |  |  | Bolivia |  |  | Brazil |  |  | Chile |  |  | Colombia |  |  | Ecuador |  |  | Guatemala |  |  |
| Cleft lip only | 74 (5) | 0.32 (1.84) | 5.41 | 101 (10) | -1.62 (1.62) | 43.56 | 1,303 (11) | -0.46 (1.73) | 15.27 | 91 (6) | -0.34 (1.64) | 15.38 | 184 (9) | -0.29 (1.63) | 10.33 | 76 (1) | -0.38 (1.73) | 14.47 | 117 (13) | 1.89 (2.38) | 8.55 |
| Cleft palate only | 160 (11) | 0.15 (1.43) | 6.88 | 68 (7) | -0.53 (1.88) | 22.06 | 2,367 (20) | -0.27 (1.62) | 12.04 | 279 (19) | -0.19 (1.29) | 6.81 | 370 (18) | -0.33 (1.65) | 13.24 | 145 (11) | -0.20 (1.58) | 10.34 | 35 (4) | 2.06 (2.61) | 8.57 |
| Cleft lip and palate | 1,200 (84) | -0.27 (1.53) | 10.67 | 831 (83) | -0.53 (1.35) | 18.77 | 8,005 (69) | -0.54 (1.65) | 15.57 | 1,090 (75) | -0.31 (1.37) | 8.07 | 1,525 (73) | -0.75 (1.77) | 19.87 | 873 (80) | -0.63 (1.82) | 20.39 | 747 (83) | 0.53 (3.03) | 25.47 |
| All clefts | 1,434 (100) | -0.20 (1.43) | 9.97 | 1,000 (100) | -0.53 (1.54) | 21.50 | 11,675 (100) | -0.47 (1.44) | 14.82 | 1,460 (100) | -0.29 (1.66) | 8.29 | 2,079 (100) | -0.63 (1.74) | 17.85 | 1,094 | -0.56 (1.79) | 18.65 | 899 (100) | 0.77 (2.99) | 22.69 |
|  | Honduras |  |  | Mexico |  |  | Nicaragua |  |  | Peru |  |  | Venezuela |  |  |  |  |  |  |  |  |
| Cleft lip only | 35 (10) | 0.51 (1.97) | 5.71 | 656 (8) | -0.82 (1.87) | 23.17 | 22 (5) | -0.16 (1.40) | 18.18 | 339 (9) | -0.26 (1.38) | 8.26 | 22 (6) | -0.24 (1.42) | 4.55 |  |  |  |  |  |  |
| Cleft palate only | 45 (12) | 0.15 (1.75) | 6.67 | 792 (10) | -0.55 (1.77) | 16.04 | 48 (12) | -0.49 (1.58) | 8.33 | 393 (11) | -0.18 (1.34) | 7.12 | 59 (16) | -0.05 (1.35) | 6.78 |  |  |  |  |  |  |
| Cleft lip and palate | 288 (78) | -0.34 (1.82) | 15.97 | 6,807 (82) | -1.03 (1.83) | 25.86 | 344 (83) | -0.63 (1.66) | 16.28 | 2,993 (80) | -0.54 (1.50) | 14.37 | 283 (78) | -0.11 (1.53) | 9.89 |  |  |  |  |  |  |
| All clefts | 368 (100) | -0.20 (1.85) | 13.86 | 8,255 (100) | -0.97 (1.84) | 24.70 | 414 (100) | -0.59 (1.64) | 15.46 | 3,725 (100) | -0.47 (1.47) | 13.05 | 364 (100) | -0.11 (1.50) | 9.07 |  |  |  |  |  |  |
| South-East Asia region |  |  |  |  |  |  |  |  |  |  |  |  |  |  |  |  |  |  |  |  |  |
|  | Bangladesh |  |  | India |  |  | Indonesia |  |  | Myanmar |  |  | Nepal |  |  | Sri Lanka |  |  | Thailand |  |  |
| Cleft lip only | 4,687 (18) | -1.60 (1.65) | 40.54 | 32,003 (15) | -1.79 (1.65) | 44.84 | 7,640 (20) | -0.83 (1.59) | 21.10 | 621 (18) | -0.83 (1.76) | 23.83 | 1,481 (19) | -1.45 (1.60) | 38.08 | 63 (4) | -1.36 (1.44) | 28.57 | 154 (10) | -1.15 (1.68) | 27.92 |
| Cleft palate only | 4,068 (16) | -1.37 (1.63) | 34.81 | 34,008 (16) | -1.33 (1.32) | 33.07 | 3,734 (10) | -0.67 (1.58) | 18.34 | 332 (9) | -1.04 (1.65) | 27.71 | 1,035 (13) | -0.87 (1.52) | 23.38 | 399 (27) | -1.62 (1.42) | 34.09 | 251 (16) | -0.89 (1.59) | 19.52 |
| Cleft lip and palate | 17,348 (66) | -1.81 (1.61) | 46.29 | 140,664 (68) | -1.82 (1.66) | 46.56 | 27,023 (70) | -0.95 (1.51) | 22.57 | 2,555 (73) | 1.01 (1.76) | 26.97 | 5,311 (68) | -1.45 (1.63) | 36.51 | 1,037 (69) | -1.56 (1.38) | 33.94 | 1,119 (73) | -1.50 (1.59) | 34.23 |
| All clefts | 26,103 (100) | -1.70 (1.63) | 43.47 | 206,675 (100) | -1.73 (1.66) | 44.07 | 38,397 (100) | -0.90 (1.54) | 21.87 | 3,508 (100) | -0.98 (1.75) | 26.48 | 7,827 (100) | -1.38 (1.62) | 35.07 | 1,499 (100) | -1.57 (1.39) | 33.76 | 1,524 (100) | -1.37 (1.62) | 31.17 |
| Western Pacific region |  |  |  |  |  |  |  |  |  |  |  |  |  |  |  |  |  |  |  |  |  |
|  | Cambodia |  |  | China |  |  | Lao PDR |  |  | Mongolia |  |  | Philippines |  |  | Viet Nam |  |  |  |  |  |
| Cleft lip only | 431 (9) | -0.99 (1.50) | 23.43 | 35,930 (25) | 0.02 (1.52) | 8.17 | 199 (22) | -0.70 (1.83) | 23.12 | 86 (8) | 0.37 (1.13) | 2.33 | 4,816 (18) | -0.97 (1.46) | 21.86 | 2,330 (23) | -0.77 (1.70) | 18.24 |  |  |  |
| Cleft palate only | 1,260 (25) | -1.14 (1.43) | 24.29 | 46,182 (32) | 0.13 (1.46) | 6.11 | 158 (18) | -0.89 (1.87) | 28.48 | 180 (17) | 0.41 (1.21) | 3.89 | 2,292 (9) | -0.81 (1.38) | 16.62 | 3,072 (30) | -0.44 (1.41) | 11.91 |  |  |  |
| Cleft lip and palate | 3,269 (66) | -1.29 (1.51) | 30.19 | 60,372 (42) | -0.11 (1.51) | 9.52 | 545 (60) | -0.98 (1.76) | 26.24 | 814 (75) | 0.04 (1.28) | 6.14 | 19,067 (73) | -1.20 (1.46) | 26.47 | 4,701 (47) | -0.66 (1.35) | 12.89 |  |  |  |
| All clefts | 4,960 (100) | -1.23 (1.49) | 28.10 | 142,484 (100) | -0.00 (1.50) | 8.08 | 902 (100) | -0.60 (1.80) | 25.94 | 1,080 (100) | 0.13 (1.27) | 5.46 | 26,175 (100) | -1.13 (1.46) | 24.76 | 10,103 (100) | -0.62 (1.46) | 13.83 |  |  |  |

